# Understanding the Incidence of Covid-19 among the police force in Maharashtra through a mixed approach

**DOI:** 10.1101/2020.06.11.20125104

**Authors:** Pravin Kokane, Priya Maurya, T Muhammad

## Abstract

**Background:** The study tries to understand the incidence of COVID-19 among police officials along with the challenges they face and their preparedness during the pandemic response in Maharashtra.

**Method:** The study analyzed the daily trends of confirmed, active, recovered, and deceased cases for Maharashtra and police professional. Ten telephonic in-depth interviews and a descriptive survey were conducted to obtain experiences of police regarding their combat against Covid-19.

**Results:** PPR (0.01 to 1.12), CRR (0 to 39.22) and CFR (0 to 1.07) have consistently increased and CRR found lower among police than the general population. The qualitative data by analyzing several indicators suggests that there is a higher individual efficacy over collective efficacy among the police force. Further, the long-time fight against Covid-19 had drained police force mentally and physically and this put them in higher risk.

**Conclusion:** Immediate priority interventions like provision of protective gears need to be provided by the government to control the risk of infection among police. Holistic support and recovery system from all stakeholders of society needed for the well-being of the police force so that they can soldier on to avert such a crisis in future.

## Introduction

A newly discovered severe acute respiratory syndrome coronavirus 2 (SARS-CoV-2) caused an infectious Coronavirus disease (COVID-19) [WHO, 2020]. It is genetically related to coronavirus responsible for the SARS outbreak of 2003, and the case fatality rate is much higher in nCOVID-2 (1). The SARS-CoV-2 disease can spread from person to person through small droplets from nose or mouth, which are spread when a person with this virus coughs or exhales. These droplets fall on the surfaces and objects around the infected person, by touching these objects and surfaces, then touching their mouth and nose, people get affected by nCOVID-19 (2). The very first case was reported in Wuhan city of China on 31^st^ December 2019. After its outbreak in Wuhan, China, the outbreak quickly spread around the world in a short span of time and become one of the severe health crises at the global level (3). In the alarming level of spread and severity of the infectious disease, the World Health Organization has declared COVID-19 as a global pandemic on 11^th^ March 2020 (4). The pandemic affected people almost all countries of the world; people belong to all socio-economic groups and races.

The nCOVID-19 is responsible for millions of infections and deaths worldwide. Till date (5th June 2020), it spreads 216 countries of the world. At the global level, 6,515,796 cases and 387,298 deaths have been reported. The USA has recorded the highest cases and deaths in the world (5). In India, the very first case was diagnosed in Kerala on 30^th^ January 2020. Earlier, the fatality rate was very low. The Indian government announced the nationwide lockdown on 24^th^ March 2020 to prevent the spread of infectious disease. India has, so far, reported 219,649 cases and 6,348 deaths with 49.45% recovery rate (including 110,960 active cases and 109,461 cured/migrated) (6).

“I really do not know where I might have been infected; I was only worried about the possibility of infecting others.” Cooke, a UK senior police officer with COVID-19 says (7). Another immigrant policeman at Bangkok international airport, after observing symptoms of fever after daily routine work visited a physician at a hospital and was diagnosed with COVID-19. He had no history of travel aboard neither any contact with confirmed COVID-19 case in Thailand. The infection is believed to have occurred unknowingly, while on duty due to coming in contact with a possible COVID-19 infected foreign passenger (8).

Historical experience suggests strongly that the speed and dread of pandemic influenza would indeed endanger internal state security (9). The incidents such as the epidemic of plague in the Indian city of Surat in 1994 which added an unprecedented level of anxiety across India, with fear and ignorance combining to freeze out even basic inter-personal sentiments of caring and civility (10), demonstrate the panic caused when populations imagine a disease out of control, and where governments are seemingly incapable of securing the safety of their citizens (11). The absenteeism across multiple sectors related to personal illness, illness in family members, fear of contagion, or public health measures to limit contact with others could also threaten the functioning of critical infrastructure and a pandemic would thus have significant implications for the basic functioning of society (12).

Paul Slovic et al. (1980) writing about risk perceptions, observed that the society appears to react more strongly to infrequent large losses of life than to frequent small losses (13). Many a conflict have emerged as police seek to enforce ‘social distancing’ measures upon communities who have been systematically disadvantaged by them. Given the measures taken by governments to control disease often produce outcomes that can threaten the very basis of functional democracy (14). Further, panic and fear are destroying the mental peace of the public with a potential to explode into irrational behaviour and social chaos, thus superseding evidence and jeopardizing the pandemic control efforts (15). To maintain public order, police personnel may be encouraged to augment their safety first and follow a systematic approach to contain the pandemic and soldier on.

Studies suggest that the greatest risk in COVID-19 is transmission to healthcare workers (16). The inadequate medical facilities and shortages of health personnel created a miserable situation in many parts of India during influenza pandemic 1918 (17). In the SARS outbreak of 2002, 21% of those affected were healthcare workers (18). Since a significant proportion of cases are related to occupational exposure, these at-risk groups which include the local police personnel should be given adequate social and mental health support (19).

Worldwide, more than half of the population stay at home to reduce transmission of disease; healthcare professionals are frontline fighter to fight COVID-19. Along with healthcare professionals, the police officials are doing the laudable job and safeguarding citizens throughout the country in the fight against this pandemic with inadequate security measures. Police officials are working in extremely challenging conditions for long hours and carry a significant burden of coronavirus. The Maharashtra police officials are ensuring strict enforcement of lockdown in the state despite hazy guidelines during this response. Enforcing lockdown on such a wide scale and ensure that Maharashtra’s 11.2 crore population stayed safe indoors, police had to respond quickly and monitored stringent measures. It is observed that constables who are frontline workers at the field were ill-informed and equipped with obsolete equipment. And this has resulted in increased infection of coronavirus among Maharashtra police. Especially, Mumbai police have accounted for the highest case of COVID-19 and causalities from other activities.

The central question, therefore, emerges concerning how the police should approach enforcement in this new, unprecedented, and insecure COVID-19 reality and what the measures government can take to ensure continuity of security and to prevent transmission of infection to the community. No such study has been done for police professionals who engaged in safeguarding citizens at any level in India. Hence, this study tries to understand the incidence of COVID-19 on police officials personally and professionally along with the challenges they had faced during the pandemic response in Maharashtra and suggest policy recommendations based on experiences and gaps in pandemic response.

## Methods

### Study design

The study was conducted among police personnel of Maharashtra. Purposely this area has been selected because COVID-19 cases are very high among police professionals. This study is based on both primary and secondary data. This study used secondary data of COVID cases from the data-sharing portal https://www.covid19india.org/. This website provides data related to COVID-19 in India and its states. Data on COVID-19 among police professionals are taken from the official twitter handle of home minister of Maharashtra https://twitter.com/AnilDeshmukhNCP). The number of COVID-19 cases categorized into 5 phases i.e., before lockdown (before 24^th^ March 2020), 1^st^ phase of lockdown (25^th^ March to 14^th^ April 2020), 2^nd^ phase of lockdown (15^th^ April to 3^rd^ May 2020,) 3^rd^ phase of lockdown (4^th^ May to 17^th^ May 2020), and 4^th^ phase of lockdown (18^th^ May to 31^st^ May 2020).

In this critical condition of infectious disease, it was not feasible to conduct a population-based survey. We selected an online data collection method for primary data. The sample size was not predetermined. We did a qualitative study through 10 telephonic interviews with seven from Mumbai police and three were from the rest part of Maharashtra to obtain the detailed descriptions of the experiences of police officials regarding the incidence of Covid-19 and lessons learned during the enforcement of lockdown. More weightage had been given to the Mumbai police because the incidence of Covid-19 cases among police personnel is high in Mumbai. All the police personnel had worked on the field since the announcement of lockdown (25th March 2020). Among this, two personnel were interviewed who tested positive in swab test of Covid-19 and three personnel recovered from corona infected were also interviewed. Purpose and objective of the study were well explained to participants. Oral consent had been taken from participants to record a telephonic interview.

Semi-structured, in-depth interviews were done with police professionals, and all interviews were audio-recorded. Secrecy has been kept by giving code numbers (R1. R2… to R10) instead of names. We adhered to standards and procedures of qualitative research reporting during the study. For quantitative data, a descriptive cross-sectional survey was conducted from 27^th^ May to 1^st^ June 2020. A close-ended questionnaire was prepared on google form, asking the participants regarding the incidence and impact of COVID-19 and associated challenges facing in this situation, and the questionnaire was shared through email, social networks (e.g., Facebook and WhatsApp). A questionnaire was circulated in a such a way that it would cover response from major cities of Maharashtra namely Mumbai, Pune, Nashik, Aurangabad, Nagpur, Amravati, Solapur, Satara, Kolhapur, Solapur etc. The questionnaire was constructed into three sections: demographic and other characteristics, collective efficacy, and individual efficacy.

### Statistical Analysis

All the analysis was performed by using Microsoft EXCEL. Frequencies and percentage distribution analyses were performed on primary data. We analyzed the trends of daily reported confirmed, active, recovered, and death cases in lockdown phases-wise in Maharashtra for the general population and the police professionals. We calculated the period prevalence rate (PPR), the case recovery rate (CRR), and the case fatality rate (CFR) phase-wise using this formula:

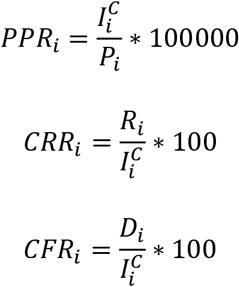

Where 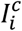 is the total COVID-19 confirmed cases in the i^th^ lockdown phase; P_i_ is the total population of the state and total police population for police professionals; R_i_ is the number of recovered in the i^th^ lockdown phase, and D_i_ is the number of deaths i^th^ lockdown phase.

## Findings

### Covid-19 pandemic in Maharashtra

In the second-most populous state of India, 580 out of every one million people have tested positive for COVID-19, which is higher than the national mark (143.14 per million). For every 100 confirmed cases, 53 cases are currently infected, 43 cases recovered from the infection and for every 100 confirmed cases, 3 have unfortunately lost their lives from the virus in the state. COVID-19 spread rapidly from a single city (Mumbai) to all over the state in a very short span of time. And this sudden increase of virus quickly overwhelmed public health and associated infrastructure in the state. **Fig 1** reflects the epidemic curve of confirmed, active, recovered, and death cases of COVID-19 in Maharashtra in five phases. Before the 1^st^ phase of lockdown, cases were very low. Cases are continuously increasing in Maharashtra. The epidemic curve also reflects what may be a mixed outbreak pattern, with early cases suggestive of a common source of foreign arrivals, and later cases suggestive of a source of unpreparedness as the virus began to be transmitted from person to person.

**Figure 1:**
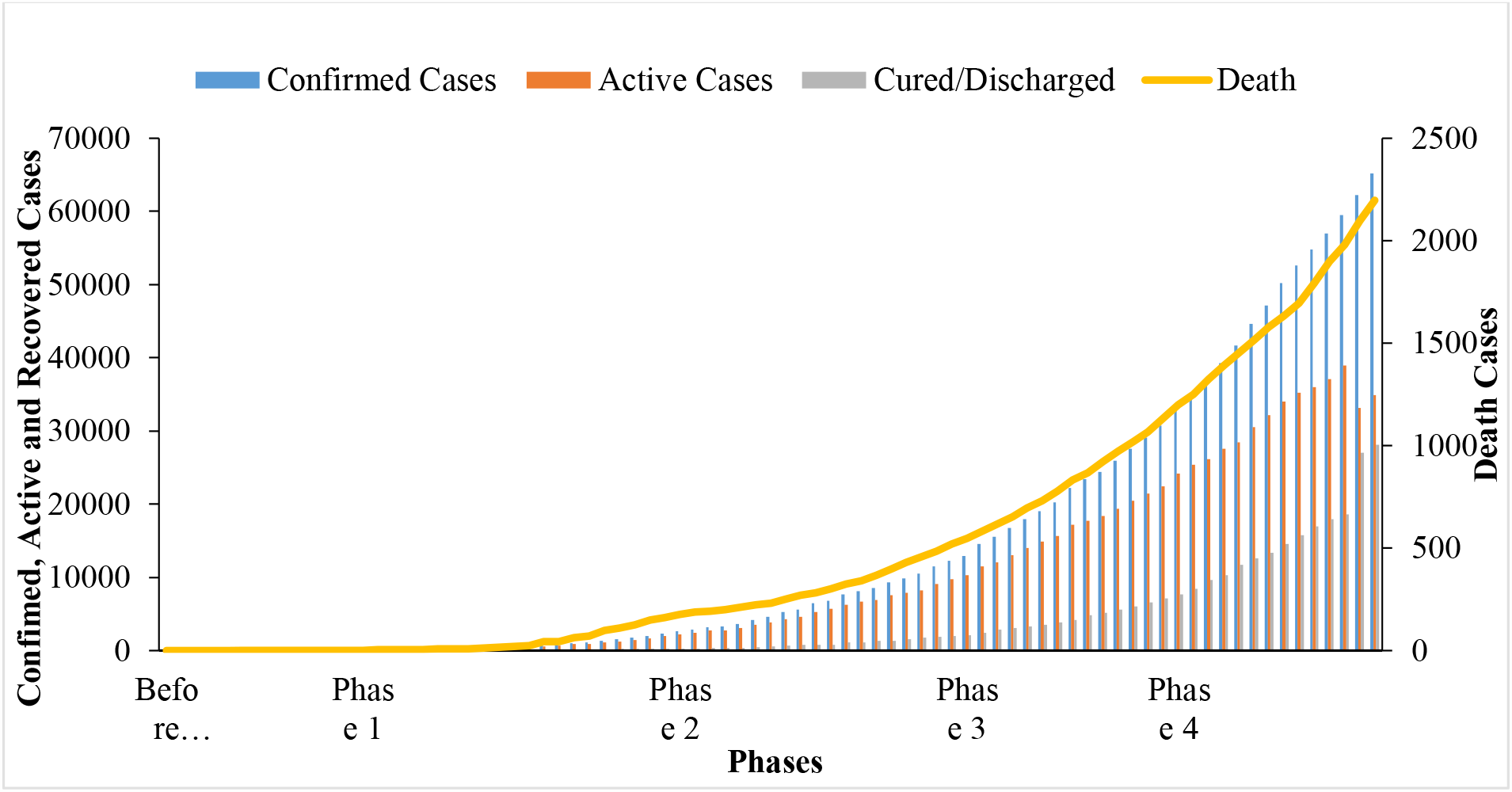
Epidemic curve of the Confirmed, Active, Cured and Death Cases of COVID-19 in Maharashtra. Source: https://www.covid19india.org/

### Police force in the frontline-

Social distancing - the key to contain pandemic by interrupting the transmission of the virus - has several dimensions which include refraining from non-essential use of public transport and avoiding large and small gatherings namely dining out, socializing and visiting other places where infections can spread easily. For this cause, the police force is in the front to fight against this pandemic. They have to manage the movement of labourers in train and buses and crowd control during a lockdown. Since the police force risked their lives on the streets to ensure the safety of citizens, it is found that 10 out of 1000 police officials tested positive from COVID-19, around 60 per cent are active cases and recovery rate is 39 percent among police professionals. The case fatality rate is lower (1.02 percent) than the whole population; this might be because of better health status and less vulnerable age group of the police force.

The police force, as one of the essential services like healthcare workers, is at the forefront of the Herculean task of enforcing the unprecedented nationwide lockdown. **Fig 2** reflects the total number of confirmed, active, and recovered cases trend. The graphs reveal that the trend in the transmission of Covid-19 among the general population in the state as well as police force are going upwards and peak number of corona cases are reported in the last two weeks of May. After analyzing the data, there were 2514 confirmed cases of COVID-19 on 31^st^ May 2020 in Maharashtra. However, the total number of deaths reported was 27. Surprisingly, there was also a significant number of 986 recovered cases till 31^st^ May 2020.

**Figure 2:**
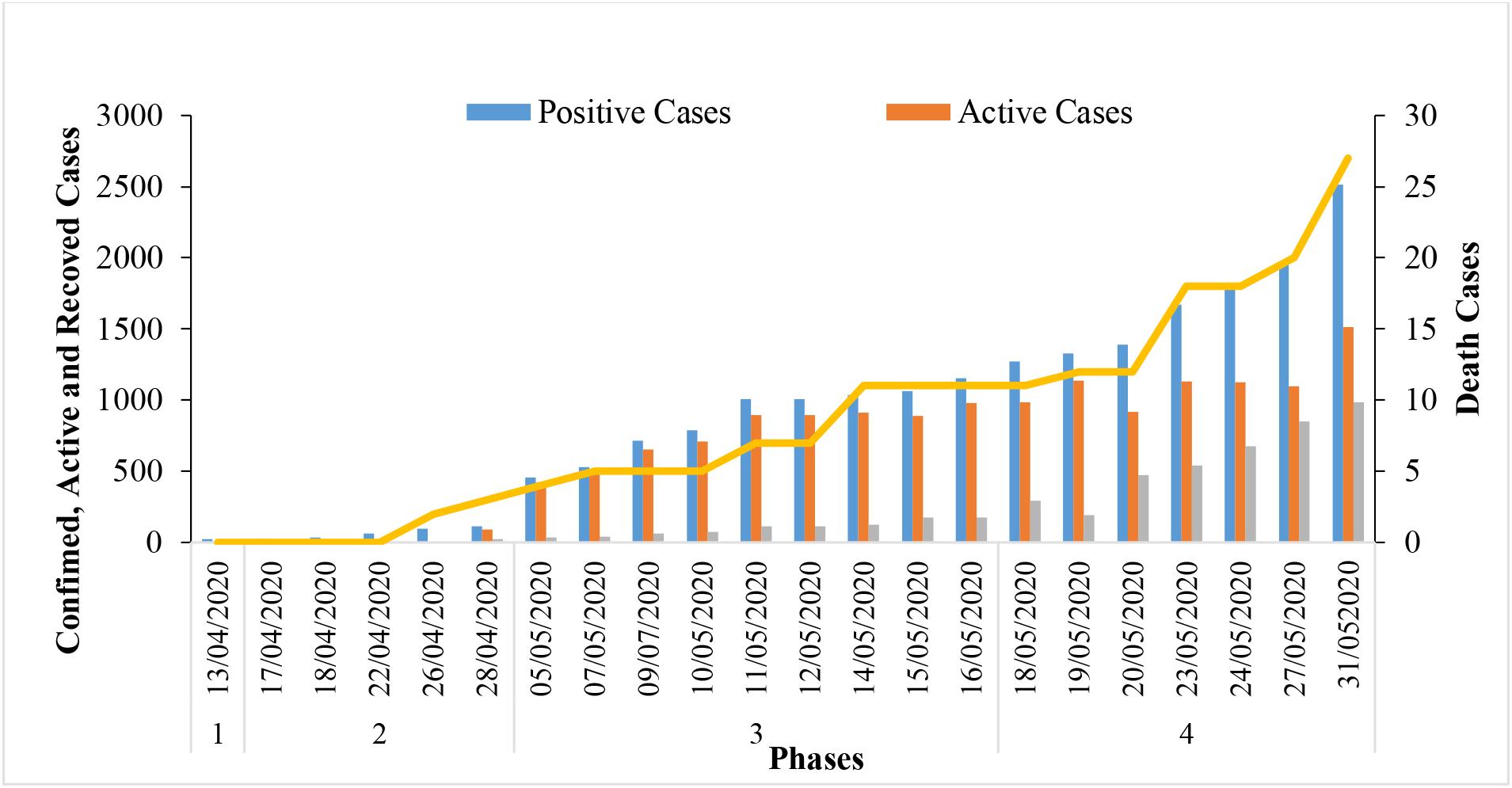
Epidemic curve of the Confirmed, Active, Cured and Death Cases of COVID-19 among Police Professionals in Maharashtra. Source: https://twitter.com/AnilDeshmukhNCP

**Fig 3** shows the PPR, CRR and CFR of COVID-19 in different phases of lockdown in Maharashtra. It is observed that before the first phase of lockdown to the fourth phase at state level PPR (0.08 to 57.99), CRR (0 to 43.09) and CFR (2.30 to 3.37) has consistently increased.

**Figure 3:**
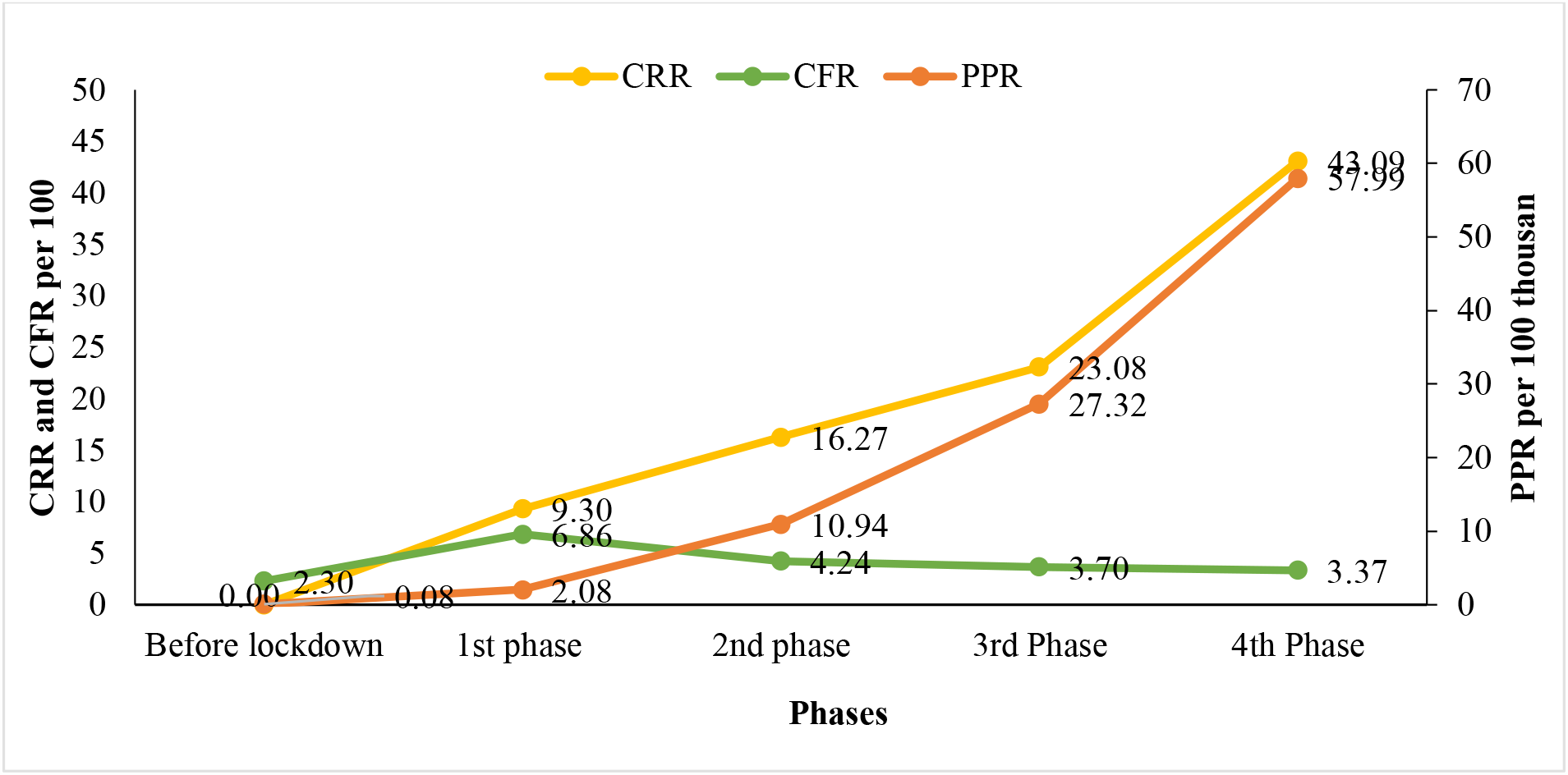
Period Prevalence Rate (PPR per 100000), Case Recovery Rate (CRR per 100) and Case Fatality Rate (CFR per 100) of COVID-19 in different phases of lockdown in Maharashtra.

**Fig 4** shows the PPR, CRR and CFR of COVID-19 in different phases of lockdown in Maharashtra Police. PPR (0.01 to 1.12), CRR (0 to 39.22) and CFR (0 to 1.07) has consistently increased. It is noticed that CRR is lower among police than the general population.

**Figure 4:**
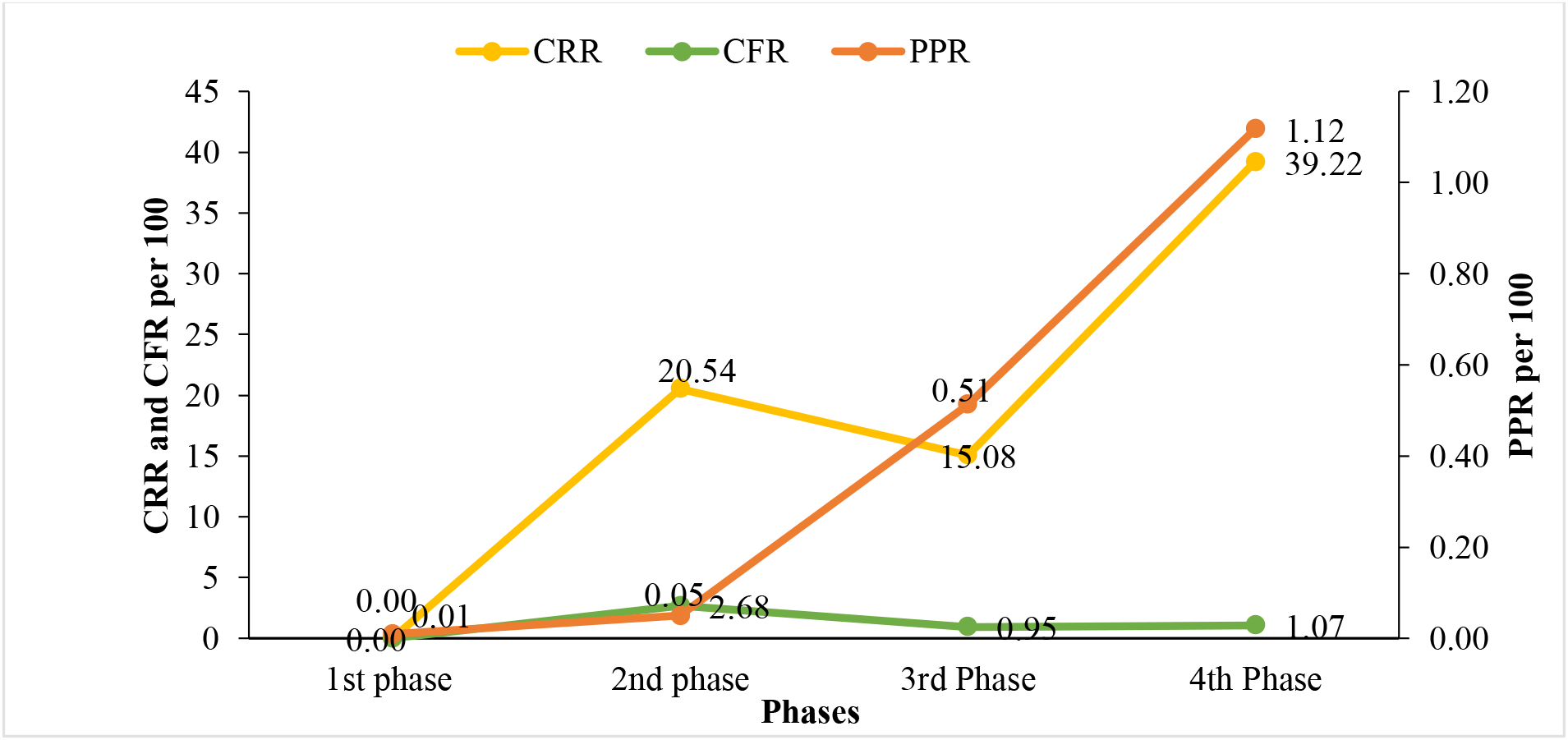
Period Prevalence Rate (PPR per 100000), Case Recovery Rate (CRR per 100) and Case Fatality Rate (CFR per 100) of COVID-19 in different phases of lockdown among Maharashtra Police.

The increase in infection cases among police force might be triggered by the states allowing migrant people to travel to homelands through public transports and private ones with special permissions even the lockdown is officially in action. Through this migration of people and in consequence of many people continued their daily life routine irrespective of government orders, the movement of people increased and resulted in increased duty time and higher exposure to the infection among police force in Maharashtra.

### Participant Characteristics of primary data-

A total of 102 respondents were included in the final analysis of primary data. The respondents were predominantly male (83.33%) and from age-group for less than 35 years (61.76%). More than half of respondents were not taking food on time, and 61% of respondents reported they did not receive any kind of supports from the local people in fighting with the virus. 12 percent of respondents exposed to COVID-19 patients during working hours **(Table 1)**.

**Table 1:** Participant Characteristics (N=102).

| **Variables** | **Frequency** | **Percent** |
| --- | --- | --- |
| **Age** |  |  |
| Less than 35 | 63 | 61.76 |
| 35-50 | 23 | 22.55 |
| More than 50 | 16 | 15.69 |
| **Gender** |  |  |
| Male | 85 | 83.33 |
| Female | 17 | 16.67 |
| **Food on Time** |  |  |
| No | 54 | 52.94 |
| Yes | 30 | 29.41 |
| Sometimes | 18 | 17.65 |
| **Drinking water** |  |  |
| Drink available at the workplace | 67 | 65.69 |
| Keep water bottle | 29 | 28.43 |
| Taking other energy drinks | 6 | 5.88 |
| **Received support or help from local people** |  |  |
| No | 62 | 60.78 |
| Yes | 14 | 13.73 |
| Sometimes | 26 | 25.49 |
| **Exposed to covid-19 patients while the duty** |  |  |
| No | 58 | 56.86 |
| Yes | 12 | 11.76 |
| Do not know | 32 | 31.37 |
| **Source of information about Covid-19** |  |  |
| WhatsApp | 49 | 48.04 |
| Other social media | 70 | 68.63 |
| TV and newspaper | 70 | 68.63 |
| Government orders | 50 | 49.02 |

**Table 2.** Observed challenges and recommended changes in practice in response to COVID-19 among the police force.

| category | Challenges | Recommendations |
| --- | --- | --- |
| Self-precautionary | Long hours of duty on the field and no time for personal health | forces should be deployed in places on a shift basis |
|  | Washing their entire uniforms properly (in detergent) themselves after going home daily | Some uniforms can be distributed and washing allowances can also be provided |
|  | Accept food or water offered to them by the local people or NGOs who are susceptible to infection | Day-to-day food and water must be provided by the department and the force would not have to depend on anyone else |
|  | No proper cleaning and disinfection guidelines and inadequate protective gears | A guidelines mechanism should be prepared, and the force must be made aware of proper hygiene and disinfected on intervals |
| Duty related | Short of maintenance of physical distancing and adhering to face-covering recommendations | Physical barriers can be installed and necessary resources such as masks, sanitizers and hand gloves etc. should be provided |
|  | Live in the slums, chawls and small flats of government quarters with shared toilets and filthy living conditions | Government must allot residences with proper sanitary environment |
|  | Lack of guidelines on detaining a suspect caught on crime and quarantining them | Training must be provided on smooth functioning of usual duties |
|  | The same number of officers travel in one vehicle as during the usual security arrangements without social distancing | vehicles from that department should have been procured immediately (in the way vehicles are procured in the election process) |
| Structural | Authority lacks an appropriate monitoring mechanism and responsible guardianship | A dedicated department can be set up to review the functioning and estimate the resource need |
|  | accustomed to dealing with every calamity with inadequate resources in the name of discipline | A separate department providing 'Logistic Support' should be set up at the state and district level to provide resources on demand |

Throughout the telephonic interviews, and the analysis of data collection from the field, a posit structural model of individual and collective efficacies (20) were examined by investigating factors associated with them. In collective efficacy, all respondents (N=10) were interviewed as they are on field duty during lockdown and responses related to preparedness to pandemic response, precautionary measures during lockdown, sanitation and hygiene facilities at workplace and support of people for enforcement of lockdown were recorded.

“After the announcement of lockdown nationwide, I had traced the travel history of immigrants under the police station of my area, he was reluctant to come to quarantine centers despite follow up from municipal corporation workers. In this situation, I had to arrest him forcefully as he was not ready to follow screening protocols. After two days, his swab test result came positive and then I realized the gravity of pandemic. This incidence put me under mental stress as I had remained under quarantine for seven days. For the first time, I worried about my family and kids” (R1). This indicates inadequate knowledge about precautionary measures needs to take for taking citizens to quarantine centers. In the first phase, security gears like gloves, masks, PPE kit were not available to police, and this is one of the factors in the spread of corona infection among them due to direct contact with people.

“Strict enforcement of lockdown led to several problems to senior citizens. As field personnel, I need to look after their problems. I have distributed food and milk packets made by local self-help group and NGO ”(R2).

“In my police area, some people turned to purchase of grocery commodities in stores, and it was urgent need to demarcate lines to space out people in front of shops and malls”(R4).

“During containment strategies, people were often neglecting government guidelines. To make them aware, I had worn a red spiked helmet of virus shape of Covid-19. This innovative way attracts the attention of local people and I succeeded highly in making them aware” (R7).

“I got a call from a local person about the unavailability of food to migrants in shelters. After inquiry, I came to know that vehicle was not available to take food from local canteen run by a self-help group. Immediately, I took my vehicle, and collected food packets from a local canteen and distributed it to migrants and elderly people” (R9). This put police to go beyond their regular duty and put extra workload during a lockdown.

“I decided to sing a song to appeal to people to stand with the nation in the fight against Covid-19. I sang a patriotic song and appealed to help police force to monitor strict implementation of lockdown”(R10).

**Figure 5** represents the response related to the collective efficacy among police professionals. More than 90 percent of police professional agreed (37.3 percent- agree and 52.9 percent-strongly agree) that infection control practices are inappropriate to avoid the spread of the virus, and 84 percent participants agreed (46.1 percent-agree & 38.2 percent strongly agree) on inadequate preparedness guidelines to fight the virus and occurring situation. About 44 percent respondents were unable to receive precautionary protective gears from authorities to protect themselves. More than half of respondents reported that sanitation and hygiene facilities at the workplace were average.

**Figure 5:**
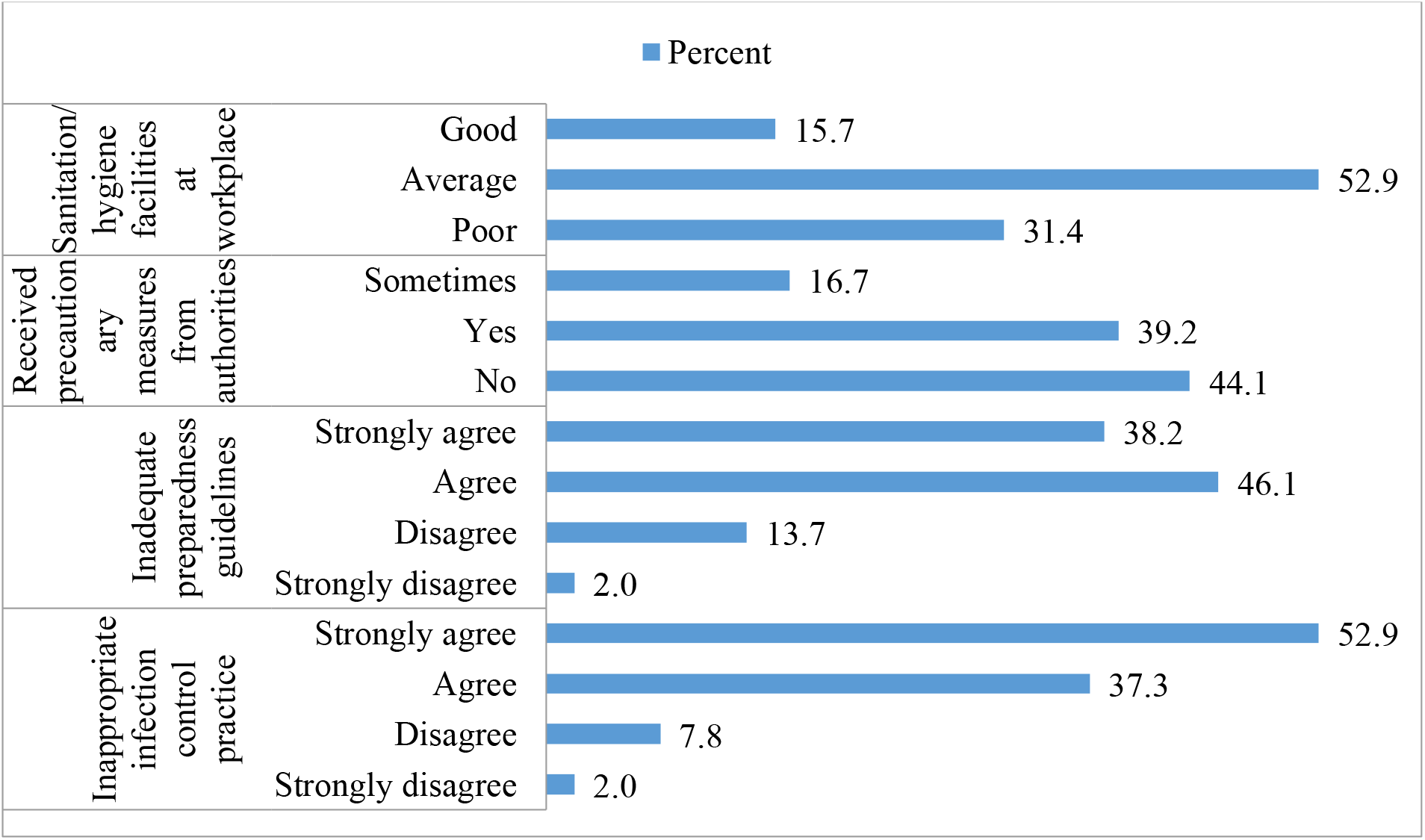
Collective efficacy towards COVD-19 among Police Professionals. *Source: calculated by authors from primary survey*

Individual efficacy theme deals with mental stress due to unavailability of protective gears on duty, risk of spreading an infection to family members and resultant fear out of it, personal health care and attention. Seven respondents were able to provide their details about the individual-efficacy theme.

“After my colleague got positive from the police station, I went under stress as I had worked and travelled along with him for barricade demarcation during day-night shifts. This made me worry about my family as we were living in a 225 square feet house near to the police station. It was difficult to maintain social distancing in such a small house as my family consists of six members”(R2).

“As I sent my family to my hometown, I was unable to clean my clothes daily. In the scorching heat of summer, I got wet completely. It was ordered to be in uniform daily, but my inability add more stress to me than relief. At the same time, wet clothes need to wash in disinfectants like sodium hypochlorite or liquid detergent. It was too difficult to get disinfectant as it went out of stock” (R3).

“Number of migrants got stranded in Mumbai and need to send to their original home with a train. I have entrusted the responsibility to look after easy travel of migrants. Some trains available were very less, and every migrant wants to go to his place. In such condition, there was bound to be in contact with migrants. Contact with migrant might be a reason for catching coronavirus infection in me as migrant labourers were staying at slum and hot spot zones of corona in Mumbai. I had to give my best in this condition to make easy movement of labourers”(R4).

“There was an inadequate supply of gloves, masks, sanitizer and PPE kit to us as priority was given to healthcare professionals. And this made me more stressed as I felt helpless in this situation. This led to constant fear, insomnia and mental disturbance in me and my colleague”(R7).

“Multiple duties and irregular shifts made difficult to take food on time. I was not able to get nutritious food, and difficult to give proper attention to health. This helplessness adds more anxiety, worry, and stress and hit badly to my immunity level. Long hours duty, seniors’ pressure to perform, no leaves, and frequent change in shifts put us in more trouble to fight against coronavirus infection.

**Figure 6** represents the response related to individual-efficacy towards COVID-19 among police professionals. Findings demonstrated that 67.7 percent of police professionals did not find time for taking care of their personal health during this pandemic and two-third respondents were taking preventive medicine of COVID-19. Only half of the respondents washed their clothes daily. One-third of police professionals were using masks, sanitizer, and gloves in working hours to protect themselves. Half of the respondents had mental disturbance due to fear of COVID-19 virus and one-third police were facing challenges of mental stress due to other reasons at the workplace.

**Figure 6:**
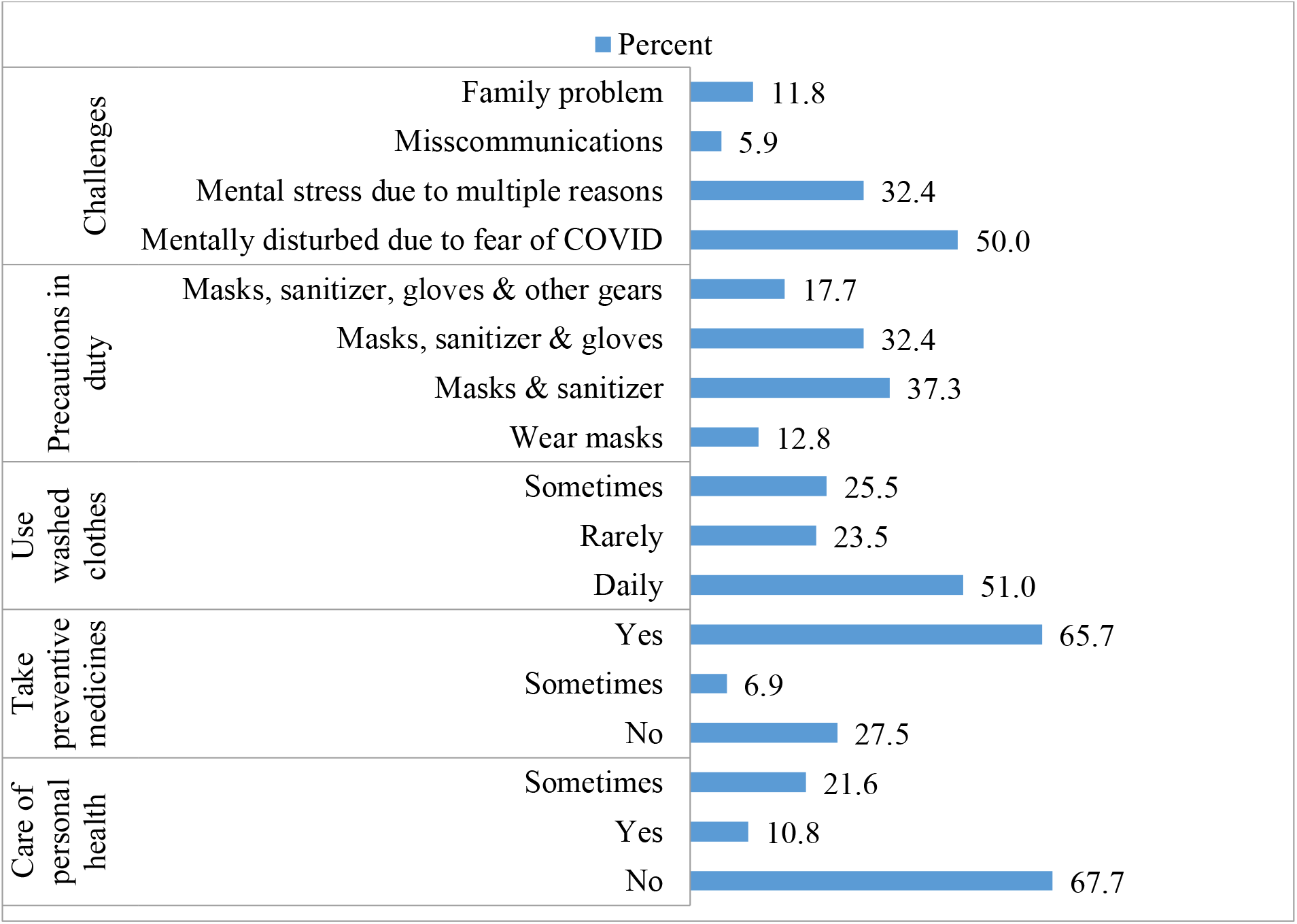
Individual-efficacy towards COVID-19 among Police Professionals. *Source: calculated by authors from primary survey*

## Discussion

To the best of our knowledge, this is the first kind of study that thoroughly assessed the incidence of Covid-19 among Maharashtra Police and their observations, suggestions regarding safety and security, and lesson learned during nationwide enforcement of lockdown in India. The study highlights the possible barriers and challenges faced by police officials in this situation. Police officials played a significant role in keeping people at home, to ensure social or physical distancing, a safe journey of inter-state migrants to their place and safeguard the whole country from the virus infection.

In this study, we identified several high-risk factors for Covid-19 infection among the police force that is rarely discussed. Our results indicate the importance of work-related problems of a police force in the local Covid-19 outbreak in Maharashtra. One novel finding is that the police force is completely dedicated to their duty and least concerned about their personal health. A finding of the study indicates that PPR and CFR have increased over the five different phases in Maharashtra as CRR is also increasing among the general population and Maharashtra police. However, the recovery rate is very low among police professionals. A previous study reported Maharashtra had observed the lowest recovery rate on an average over lockdown period (21). In the fourth phase of the lockdown number of confirmed cases among police increased sharply. Similar findings from a follow-up study reported that police officers are among the leading high-risk occupations in the late transmission period (22). In an ongoing pandemic situation and unpredictable risks, police officials also worried about their families and have mental pressure. However, still, they are fighting against the virus, took up their responsibilities, concentrated on their duties even without receiving proper safety kits. They showed a spirit of unity and professional dedication towards their work in this hard time.

Results demonstrate that 50 percent of police got information about the virus from social media (ex. WhatsApp and Facebook). Findings are consistent with other studies, as Bhagayathula et al. reported that 60% of HCPs use social media to seek information regarding COVID-19 (23). Inadequate knowledge is not the only risk factor attributed to infection; occupational exposure is also a cause of the higher risk of infection (24). This study reported that majority of the respondents agreed that infection control practices and preparedness guidelines are inappropriate and inadequate, and they did not receive precautious measures, while they are also frontline workers in fighting the pandemic. A study reported that healthcare professionals are more aware of information regarding the infectious disease, self-hygiene and to protect themselves from the virus compared to other occupations (25). It is found that only half of the respondents were using daily washed clothes, and most of them taking preventive medicine to protect themselves because they do not have proper safety gear provided by authorities. Further, a systematic review done by Rajkumar, R.P. (2020) reported that 28% of respondents have symptoms of anxiety and depression in the general population during this period (26) while in our study, 50% of police personnel are mentally disturbed due to fear of COVID-19 and faces challenges at the workplace.

Police personnel did not have a system in place to determine if the people they were dealing with were affected. Even in the absence of such an arrangement, they began to carry out their duties. But in a short time, the fact that many officers and staff on duty in the department were infected with the coronavirus came to light and this has been confirmed with an increase in corona positive cases (more than 2500 cases) in subsequent lockdown phases. A study on healthcare professionals reported that workforce safety should be on high priority to reduce uncertainty and fear (27).

This study suggests immediate priority interventions to prevent spread of coronavirus among police. At workplace, there is need to provide clean drinking water (to avoid dehydration due to heat) and handwashing facilities. Police personnel were not exempted from their intended duties i.e. prevention of crimes. Security gears (gloves, masks, face shields and PPE kit) should be given to arrest of accused as his/her corona status is unknown to police. A separate department providing logistics support should be set up at state and district level which will take care of travel, stay, food and sanitation facilities.

## Conclusion

During enforcement of lockdown, police professional went through high risk of morbidity and mortality. Long hours of duty, multiple shifts and inadequate security gears had drained police force mentally and physically. Further studies are required to notify the effectiveness of immediate priority interventions. There is need of holistic support and recovery system for the well-being of police personnel so that efficiency and preparedness of police force will be utilized effectively in future for such type of pandemic crisis.

### Limitations of Study

As far as limitations of the study are concerned, all participants for the qualitative study were interviewed telephonically as it was difficult to create rapport with participants on the telephone. Questionnaire created through electronic forms consist of closed-ended questions only, lack of open-ended questions underreported rapidly changing evidence of risk factors of police. A sample size of 102 seems reasonable to analyze experiences and lessons learnt during this pandemic.

## Data Availability

All data is available at government site and official site of Government of Maharashtra and Government of India, Data is find at this site- https://arogya.maharashtra.gov.in/1035/%e0%a4%ae%e0%a5%81%e0%a4%96%e0%a5%8d%e0%a4%af-%e0%a4%aa%e0%a5%83%e0%a4%b7%e0%a5%8d%e0%a4%a0 snd https://www.mohfw.gov.in/.

https://www.mohfw.gov.in/

## Contributors

All authors had access and involvement in the collection of data and take accuracy and responsibility for the integrity of data and its analysis. PK had conceptualized and designed the study.PM and MT contributed equally and shared the second authorship.PK contributed equally and shared the first authorship.PK collected the data, and translate it into English, and PM and MT analyzed and interpreted the data. PK, PM, and MT wrote an original draft of the manuscript. All authors contributed to reviewing and editing of paper equally.

## Acknowledgement

We deeply acknowledge support of all police personnel during interview and data collection. Special mention goes to Mr. Ashok Sabale for their support and guidance during this work.

